# Can a knee sleeve influence ground reaction forces and knee joint power during a step-down hop in participants following ACL reconstruction? An explanatory analysis

**DOI:** 10.1101/2022.08.17.22278057

**Authors:** Gisela Sole, Todd Pataky, Niels Hammer, Peter Lamb

## Abstract

**Purpose:** Elastic knee sleeves are often worn following anterior cruciate ligament reconstruction but mechanisms underlying observed changes in movement patterns are still unclear. The aim of this study was to determine the immediate and 6-week effects of wearing a knee sleeve on ground reaction forces (GRF) and knee joint power during a step-down hop task.

**Methods:** Using a cross-over design, we estimated GRF and knee kinematics and kinetics during a step-down hop for 30 participants (age 26.1 [SD 6.7] years, 14 women) following ACL reconstruction (median 16 months post-surgery) with and without wearing a knee sleeve. In a subsequent randomised clinical trial, participants in the ‘Sleeve Group’ (n=9) then wore the sleeve for 6 weeks at least 1 hour daily, while a ‘Control Group’ (n=9) did not wear the sleeve. Statistical parametric mapping (SPM) was used to compare (1) GRF trajectories in the three planes as well as knee joint power between three conditions at baseline (uninjured side, unsleeved injured and sleeved injured side); (2) within-participant changes for GRF and knee joint power trajectories from baseline to follow-up between groups. We also compared discrete peak GRFs and power, rate of (vertical) force development, and mean knee joint power in the first 5% of stance phase.

**Results:** GRF did not differ for the (unsleeved) injured compared to the uninjured sides based on SPM analysis. Discrete variables showed lower peak anterior (propulsive) GRF for the injured side, and lower peak eccentric and concentric power, and mean power in the first 5% of stance. When wearing the sleeve on the injured side, mean power in the first 5% of stance increased significantly [mean difference (95% CIs) 1.3 (0.6, 2.0) N/BW*ht] from a concentric to an eccentric power when wearing the knee sleeve. After six weeks, the direction of change for vertical GRF differed between the groups: while the Control Group had slightly decreased forces, the Sleeve Group presented increased forces.

**Conclusions:** Increased knee power in the first 5% of landing when wearing the knee sleeve, combined with greater knee flexion, may indicate a protective response for ACL ruptures, most commonly occurring during that early phase of landing. The directional change of increased vertical GRF for the Sleeve group, combined with shorter stance duration at follow-up, may indicate enhanced performance when being prescribed such sleeve.

## Background

Rupture of the anterior cruciate ligament (ACL) is a debilitating knee injury with potentially devastating short-term and long-term consequences. Rehabilitation following ACL reconstruction includes individualised progressive exercise prescription to improve range of motion, muscle strength, sensori-motor control and sports- and work-specific skills, as well as physical fitness [1]. Strategies are also included to address potential psychosocial factors, such as fear of re-injury, and to improve knee-related confidence and self-efficacy for return to physical activity [2–4]. Such strategies may include prescription of wearing a knee sleeve, or people with ACL reconstruction may intuitively use them [2, 5]. We have shown that individuals with ACL reconstruction may have immediate improved jump-related performance when wearing a knee sleeve [6]. Besides focussing on the distance or height of jumping, considering movement patterns during landing are also important [7, 8]. In our initial analysis of movement patterns during a step-down hop, we found that participants with ACL reconstruction landed with greater knee flexion when wearing a knee sleeve [9]. Wearing a sleeve for at least one hour over a 6-week period resulted in no differences in knee flexion and moments compared to participants who did not wear the sleeve, but those with the sleeve jumped faster, evidenced with shorter stance duration [9]. Wearing a knee sleeve may influence sensorimotor control [10–12], however, the mechanisms whereby a knee sleeve might improve jump distance or enhance knee flexion during jump landing are unclear.

Jump-landing strategies have received substantial attention as a risk factor for ACL rupture and as outcomes following such injury [13, 14]. Current understanding is that increased impact, reflected by higher vertical ground reaction forces (GRF), increased posterior GRF [15, 16], and higher rate of force development (RFD) may increase risk for incurring an ACL injury [17]. In uninjured athletes, higher vertical GRF during jump landing appear to be associated with decreased hip, knee and ankle flexion angles, thus a ‘stiffer’ leg during landing [18]. Such stiffer landing patterns are associated with increased risk of subsequent injuries in current uninjured participants [19]. Jump landing training can increase knee flexion on landing [14] and reduce vertical and posterior GRF and RFD [17]. Following ACL injury and ACL reconstruction, the response of GRF is less clear. Vertical GRFs are likely to be lower for the injured than the contralateral uninjured side post-reconstruction [20, 21], however such change may be time- and task-dependent. For maximum single-leg hop, a systematic review found moderate evidence for no difference between ACL-injured and contralateral sides for vertical GRF [7]. Pietrosimone et al. [22] showed that within the first 12 months following ACL reconstruction, peak vertical GRF are likely to be lower compared to the contralateral uninjured sides while walking but at mid-stance, the GRF may be higher. In contrast, in the phase from 12 to 24 months post-ACL, the peak vertical GRF are likely to be higher compared to the contralateral sides [22]. The GRF of the injured side relative to the contralateral uninjured sides may thus change over the recovery period. The desired direction for GRF change is thus uncertain post-ACL reconstruction.

During jump landing, power is absorbed by the lower extremity, and during take-off, power is generated. Lower knee range of motion decreases the range over which force can be generated, potentially leading to lower peak knee moments and knee power [7, 23]. Following ACL reconstruction, it is likely that knee power is reduced, both during absorption and during take-off [7, 23].

Our project for exploring the effects of wearing a knee sleeve during the step-down hop provides opportunity to explore the influence of wearing a knee sleeve on the ground reaction forces and on knee power. The aim of this study was to determine the (1) immediate effects and (2) 6-week effects of wearing a knee sleeve on GRF and on knee power in participants who had undergone an ACL reconstruction in the previous five years.

Our primary hypothesis is that peak vertical GRF and the rate of force development will be lower in the injured side compared to the uninjured side during the unsleeved conditions. The secondary hypothesis is that wearing the sleeve will increase the peak vertical GRF and rate of force development during the absorption phase of the injured side. Similarly, we hypothesise that, for the 6-week effects, the Sleeve Group will have larger changes in the vertical GRF and for rate of force development than the Control Group.

## Methods

This is the third paper in a sequence of papers stemming from a single, multi-year study exploring the influence of wearing a knee sleeve for people with ACLR. This paper differs from the previous papers [6, 9] in that it considers GRF and knee power. We recruited participants from August 2018 to September 2020 and the follow-up data collection was completed in October 2020. While this paper reports an additional analysis, there are no on-going or related trials for the intervention. The data had been collected during two sessions (baseline and six-week follow-up) in the School of Physiotherapy Human Movement laboratory of the University of Otago, and via REDCap (Research Electronic Data Capture). The Health and Disability Ethics Committee (of New Zealand) granted ethical approval for the study. We follow CONSORT reporting guidelines [24]. We repeat the data collection methods here for completeness of this report.

### Trial design and blinding

The study had two linked parts and all participants were involved in both parts. Part 1 consisted of a cross-over laboratory-based study, to examine immediate effects of the wearing of the knee sleeve on single-leg hop distance [6] and knee mechanics during a single-leg step-down hop task [9]. Part 2 entailed a parallel two-armed, assessor-blinded randomised controlled trial (RCT) to determine the effects of wearing the knee sleeve over a 6-week period on self-reported knee function and physical performance measures.

## Participants

### Recruitment

We recruited participants via community advertising and the research participant recruitment agency TrialFacts (https://trialfacts.com/). Volunteers completed a questionnaire (also serving as screening for eligibility) via REDCap prior to attending the first laboratory session. The questionnaire included demographics, injury and surgery history, the International Knee Documentation Committee Subjective Knee Form (IKDC-SKF) [25] and the Tegner activity scale [26]. The Tegner scale categorises sports and physical activity in terms of the level of knee-related loading where ‘0’ indicates ‘sick leave or disability due to a knee injury’ and ‘10’ indicates ‘competitive soccer or rugby at national or international elite level’.

### Inclusion criteria

We recruited men and women, aged 18-40 years, who underwent ACL reconstruction within 6 months to 5 years previously. We specifically sought individuals who had not yet reached full functional level, defined for the purpose of this study by a score between 40 to 80/100 on the IKDC-SKF [25, 27, 28].

### Exclusion criteria

Participants were excluded if they had undergone a revision ACL reconstruction of the same knee (due to re-injury), or a previous ACL reconstruction of the opposite knee; self-reported any other lower limb, pelvic or low back musculoskeletal injuries or disorders that required medical care over the past 6 months; had known systemic, neurological or cardiovascular disorders; or had a body mass index (BMI) greater than 30 kg/m^2^. Participants found to have an IKDC-SKF score less than 40 (due to potential safety risk during the laboratory-based tasks) or greater than 80/100 (as use of a sleeve would clinically be less likely to add benefit) were excluded.

## Procedures

### Randomisation

Participants were individually randomised twice (once for the cross-over trial, and once for the RCT) with equal numbers in each group for both allocations. Block randomisation (in groups of 8 participants) was undertaken sequentially by a research officer using an electronic random number generator prior to participants being entered into the study. Each group was stratified by sex. The research officer informed the researcher responsible for the laboratory data collection of the order for the conditions for the cross-over trial, and the group allocation (for the RCT) via email prior to the start of the individual participant’s first laboratory session.

Eligibility to be included was confirmed and participants provided written informed consent at the start of the first session. Participants were asked to be dressed in a singlet, a pair of shorts and their own sport shoes. Body mass and height were measured during the baseline session.

#### Part 1: Laboratory cross-over trial – immediate effects

Participants undertook two hopping tasks; a maximum horizontal single leg hop and a stub-maximum step-down hop. Participants practised the hopping tasks at sub-maximal distance with the uninjured and injured sides until they were confident with performing them as part of familiarisation and warm-up. They performed the maximum horizontal hop prior to undertaking the step-down-hop.

#### Part 2: Randomised clinical trial

Participants were informed of their group allocation for the RCT on completion of the first laboratory session. Following the 6-week period, all participants were asked to return to the laboratory to repeat the above assessments, repeating the hopping tasks (without wearing the knee sleeve).

### Intervention

The intervention entailed use of the commercially available GenuTrain (Bauerfeind^®^ AG, Zeulenroda-Triebes, Germany), a CE-certified medical device. For Part 1 (cross-over trial), all participants performed the step-down hop with and without the sleeve. For Part 2 (RCT), participants of the ‘Sleeve Group’ (intervention) were instructed to wear the knee sleeve while performing their rehabilitative exercises, physical activity and sports, with a minimum of 1 hour per day for the 6-week period; the control group were not provided with a sleeve during this period.

## Outcomes

<u>Step-down hop</u>: Three-dimensional motion analysis was performed for the step-down hop with 11 infra-red Eagle-500RT cameras (Motion Analysis Corporation, Santa Rosa, CA, USA), sampling at 120 Hz, and Cortex 4.4 software (Motion Analysis Corporation, Santa Rosa, CA, USA). This was synchronized with a floor-mounted tri-axial force plate (OR6-5 AMTI Inc., Newton, MA, USA), sampling at 2,400 Hz. Cortex 5.5 was used to track and label the markers, and the biomechanical model, kinematic (joint angles) and kinetic (moments) variables were calculated using Visual3D Professional v6 (C-Motion, Inc., Germantown, MD, USA). Here we report the procedures only for the force plate data.

The participants were asked to stand on a 30-cm box, placed 15 cm from the force plate, and performed a step-down hop (adapted from E Kristianslund and T Krosshaug [29]) onto the force plate: the participants were asked to step off the box with either the injured or the uninjured leg onto the force plate, then hop forward off the plate as fast as possible. The distance of that hop was defined as 60-70% of the maximum horizontal jump length. They performed the step-down hop with the uninjured side first, then the injured side under the (1) the ‘sleeved’ condition (experimental, wearing the sleeve) and (2) the ‘unsleeved’ condition (control, no sleeve), ordered by randomisation. A 5-minute walk between the conditions provided a standardised run-in to the second condition to minimise carryover effects.

## Data processing

GRF data from the stance phase of the hop is of interest; the start and end of the stance phase were defined by the vertical component of the GRF exceeding and returning below 20 N, respectively. Based on the SPM analysis the following discrete variables were extracted: rate of force development, mean joint power during the first 5% of stance, peak eccentric joint power and peak concentric joint power. The averages of five trials for each limb (injured versus uninjured) and condition (sleeved and unsleeved) for each participant along with descriptive variables were calculated.

The *x*, *y*, and *z* components of the ground reaction force data during stance were analysed for immediate effects (Part 1) and 6-week effects (Part 2). Force data were time-normalised to 1001 data points for each participant and condition (baseline) or session (follow-up). The mean time of all first and second vertical GRF peaks (*F_z_* peaks), relative to the mean length of all trials, were used to time-align the respective *F_z_* peaks for each trial. Therefore, data were time-normalised in three phases to ensure comparison of equivalent events in the movement. For trials with only one *F_z_* peak, standard time-normalisation to 1001 frames was performed.

Joint power was analysed as a follow-up to the GRF presented in this report and the kinetic, kinematic and temporal variables in previous reports. We investigated joint power in the sagittal plane as *JP*(*t*) = *M_x_* (*t*) · *ω_x_* (*t*), where *JP* is joint power at each time *t*, *M_x_* is the knee flexion-extension moment (normalised by body weight and height) and *ω_x_* is knee flexion-extension angular velocity. By convention a knee (external) flexion moment is positive, and a flex*ing* knee has a positive angular velocity, thus positive joint power indicates a net eccentric muscle contraction at the joint and negative joint power indicates a concentric contraction.

## Statistical analysis

For our primary analysis, we analysed GRF and knee joint power using Statistical Parametric Mapping (SPM, http://spm1d.org/; Pataky, 2012) [30]. Mean trajectories of five trials for each participant, limb and condition (Part 1) and each session (Part 2) were computed using MATLAB R2022a (The MathWorks Inc., Natick, MA, USA). A secondary analysis of discrete variables followed. The data set can be found on Zenedo [31].

### Ground reaction forces

We determined immediate effects by two-way comparisons across the three combinations of sleeved injured leg, unsleeved injured leg, uninjured leg. The SPM time-continuous Hotellings *T^2^* test was performed on the GRF components within each baseline condition [32, 33]. SPM allows comparison of the entire GRF trajectory, rather than a pre-selected discrete variable, which helps to control both Type 1 and Type 2 error rates [32].

Six-week effects were determined by calculating the mean, time-normalised GRF curves for each component, as with the immediate effects, and subtracting the baseline from the follow-up session, leaving ‘difference’ trajectories. We performed a time-continuous Hotellings *T^2^* test on the three-dimensional force difference trajectories comparing Sleeve and Control groups [32, 33]. For both immediate and 6-week analyses, significant effects were analysed with *post-hoc* time-continuous *t*-tests to determine which conditions differed.

A conservative Bonferroni threshold of 0.017 was adopted to correct for multiple comparisons across the three GRF components.

### Knee joint power

Sagittal plane joint power is a one-dimensional, time-continuous variable so immediate effects were determined with multiple time-continuous paired *t*-tests. A Bonferroni threshold of 0.017 was adopted to correct for multiple comparisons across the three conditions. Six-week effects were determined by time-continuous independent t-tests on the joint power difference trajectories. SPM calculations were performed using the spm1d package version M.0.4.8 (spm1dmatlab: One-Dimensional Statistical Parametric Mapping in MATLAB. https://github.com/0todd0000/spm1dmatlab, T Pataky, 2019).

### Secondary analysis: Discrete variables

Post hoc analyses were performed for the pre-defined GRF and knee joint power discrete variables, and those that were deemed to be of interest from the SPM analysis. Time-based variables included the Rate of Force Development (RFD). The RFD reflects the speed at which *F_z_* increases from initial contact to the first *F_z_* peak. We defined the RFD as the first *F_z_* peak divided by the time duration from landing force to the first *F_z_* peak [34]. For trials with a single *F_z_* peak a time duration of 100 ms was used.

To investigate the immediate effects of wearing a knee sleeve, we used one-way repeated measures ANOVAs to compare three conditions at the baseline: (a) uninjured side, unsleeved to (b) injured side, unsleeved, and (c) injured side, sleeved. Sex (male/women) and time since ACL reconstruction (in months) were entered as co-variates. If Mauchley’s test for sphericity was significant (p≤ 0.05), the Greenhouse-Geisser correction was used. Post-hoc pairwise analyses using paired *t*-tests and a Bonferroni correction across the three pairwise tests assessed between-condition effects.

Individual change scores from baseline to follow-up were calculated for the dependent variables of the GRF and knee joint power. Due to low sample size (n=9 per group) the change scores were compared between the intervention and the control groups using Mann-Whitney U tests for each outcome. The alpha level were set at p≤0.05. These analyses and those of demographic data were performed with SPSS Version 28.0.1.0 (IBM Corp, Armonk, NY).

## Results

We assessed 34 participants at baseline, but data for four participants were excluded from this analysis due to technical issues. Two participants of the Sleeve Group withdrew from the study following baseline assessment due to knee re-injuries, unrelated to use of the knee sleeve (Fig 1). Eight participants were lost to follow-up due to the COVID-19 lockdown in New Zealand, March/April 2020. Twenty-four participants completed the follow-up laboratory session. Data from six participants were excluded due to technical difficulties, resulting in data being analysed for nine participants in each group for Part 2 (RCT). Demographic data of the participants are provided in Table 1.

**Figure 1.**
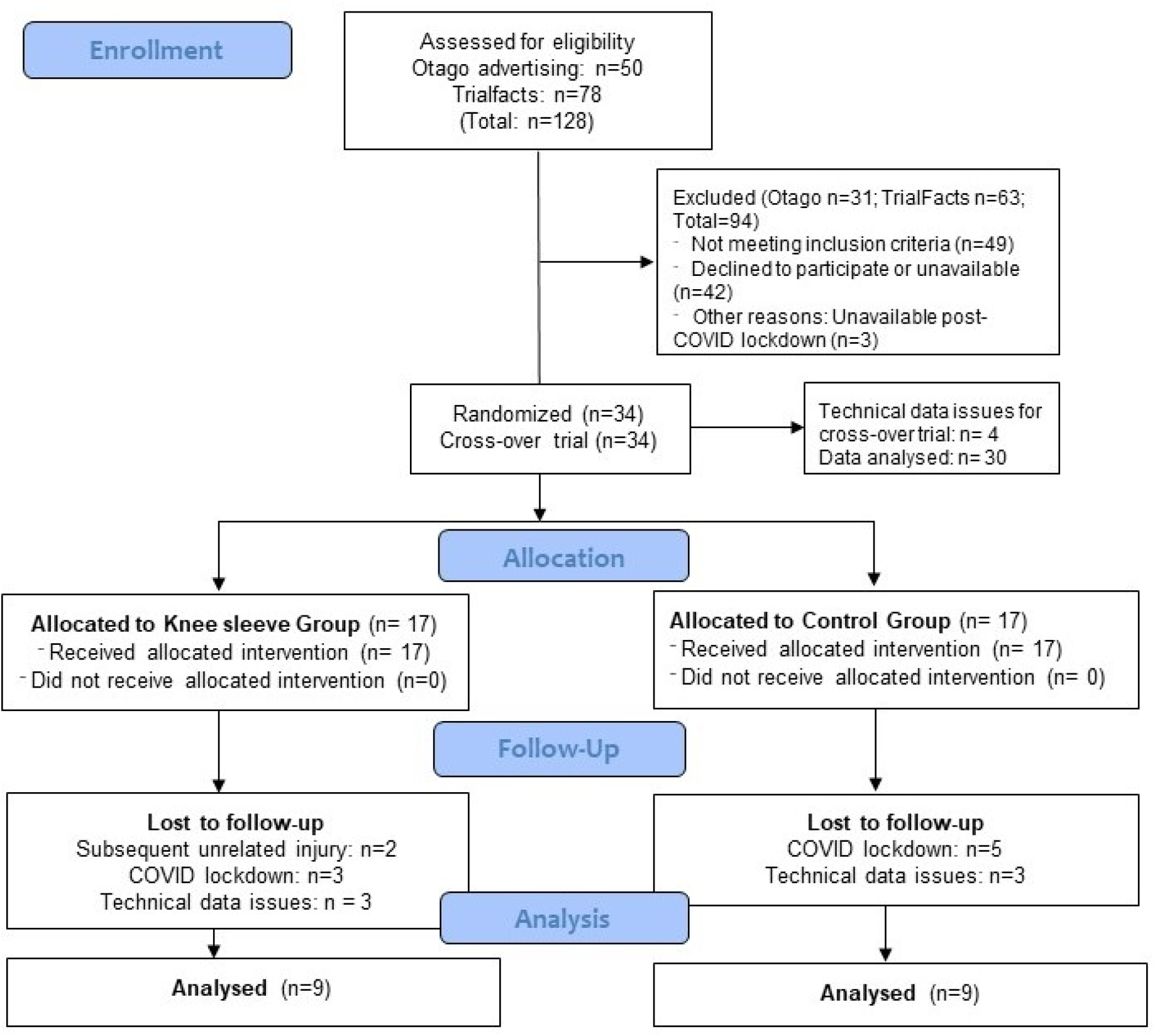
CONSORT flowchart of participant recruitment, allocation and follow-up.

**Table 1.** Demographic data (n = 30)

|  | All | Men | Women |
| --- | --- | --- | --- |
| Men/Women n (%) | 30 | 16 (53) | 14 (47) |
| Age (years) | 26.1 (6.7) | 25.5 (5.7) | 27.3 (7.6) |
| Mass (kg) | 75.9 (11.3) | 78.5 (12.2) | 76.1 (11.8) |
| Height (m) | 1.72 (0.1) | 1.76 (0.07) | 1.67 (0.08) |
| Body mass index (kg.m<sup>-2</sup>) | 25.7 (3.1) | 25.1 (2.7) | 27.1 (3.3) |
| Reconstruction: Hamstring/patella<br>tendon grafts n (%) | 14 (47)/16 (53) | 9 (56)/7<br>(44) | 5 (36)/9 (64) |
| Meniscal repair: no/yes n (%) | 22 (73)/8 (27) | 13 (81)/3<br>(19) | 9 (64)/5 (36) |
| Time since ACL injury (months) | 21 (9 – 108) | 21 (9 – 55) | 25 (12 – 108) |
| Time since surgery (months) | 16 (6 – 53) | 17 (6 – 44) | 16 (7 – 53) |
| Time from ACL injury to surgery<br>(months) | 6 (1 – 89) | 6 (1 – 11) | 8 (1 – 89) |
| Tegner activity scale: Preinjury<br>(median, range) | 8 (3 – 10) | 9 (3 – 10) | 7 (6 – 10) |
| Tegner activity scale: Baseline<br>(median, range) | 5 (2 – 9) | 5 (2 – 9) | 4 (2 – 9) |
| IKDC-SKF Baseline | 66.8 (9.8) | 67.2 (10.3) | 67.8 (8.7) |
| IKDC-SKF Follow-up | 73.7 (11.5) | 71.3 (13.2) | 76.5 (8.9) |

Figures are numbers (Frequency), Mean (SD) or Medians (minimum – maximum)
IKDC-SKF: International Knee Documentation Committee Subjective Knee Form

### Part 1: Immediate effects

#### Ground reaction forces

The SPM analysis found no statistical differences between any GRF components for any of the baseline conditions (Fig 2 to 4).

**Figure 2:**
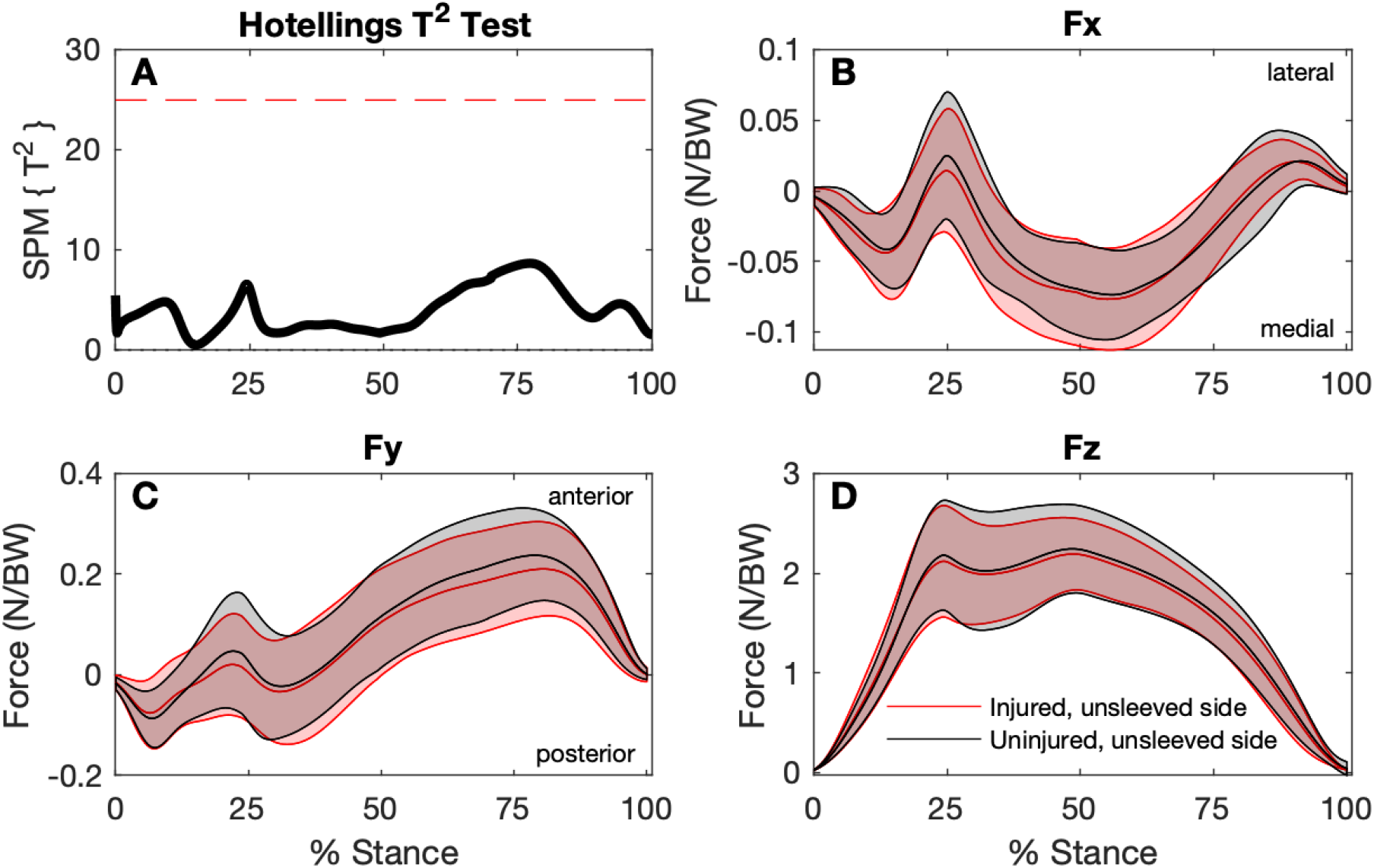
Comparison of ground reaction forces during the stance phase of the step-down hop for the injured (unsleeved) sides to the uninjured contralateral sides (n=30). A: SPM Hotellings T^2^ test trajectory, dashed red line indicates adjusted significance criterion; B to D: x (lateral/medial), y (anterior/posterior), and z (vertical) component curves (mean and ±1 standard deviation bands), respectively.

**Figure 3:**
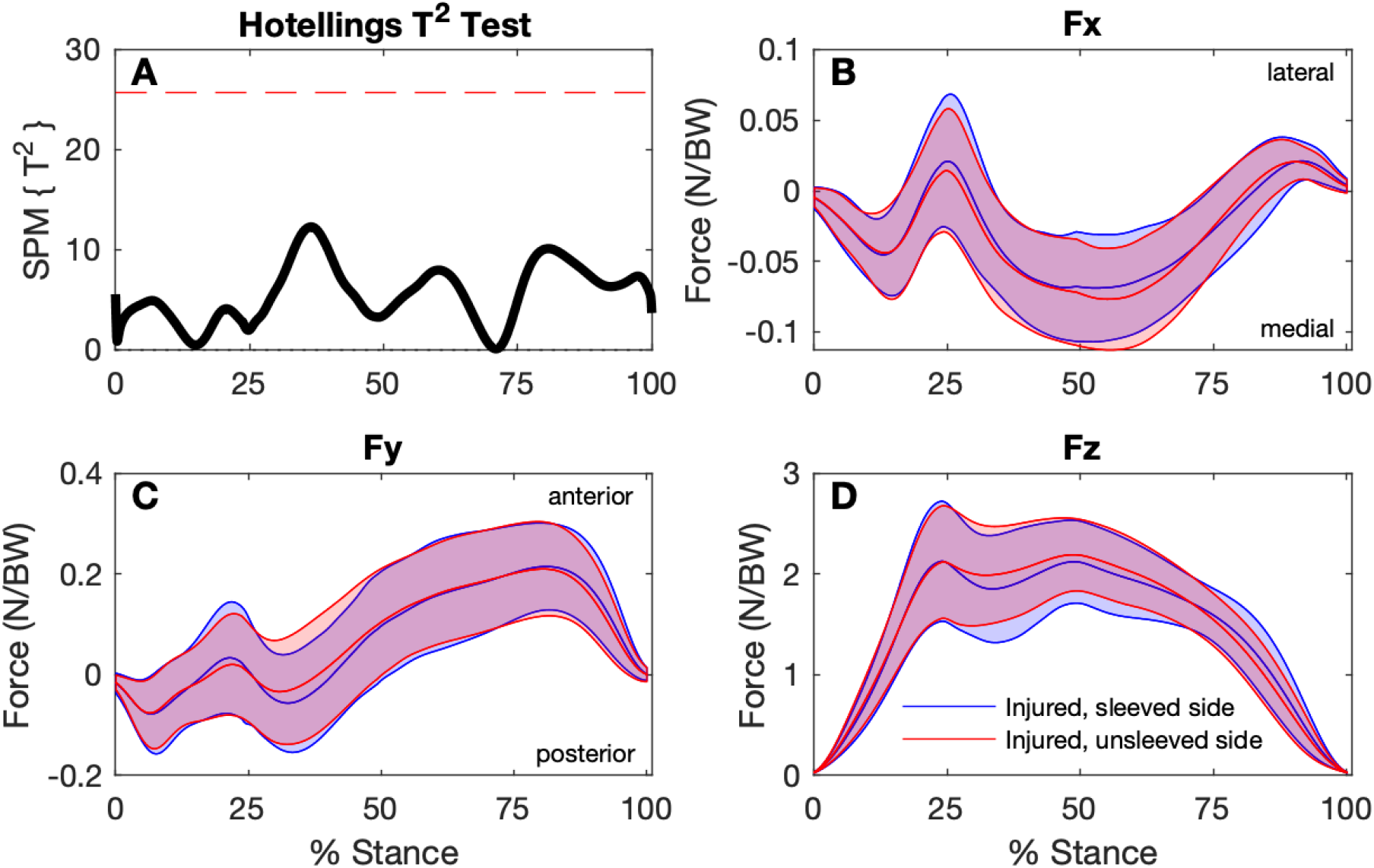
Comparison of ground reaction forces during the stance phase of the step-down hop for the sleeved and unsleeved conditions for the ACL injured sides (n=30). A: SPM Hotellings T^2^ test trajectory, dashed red line indicates adjusted significance criterion; B to D: x (lateral/medial), y (anterior/posterior), and z (vertical) component curves (mean and ±1 standard deviation bands), respectively.

**Figure 4:**
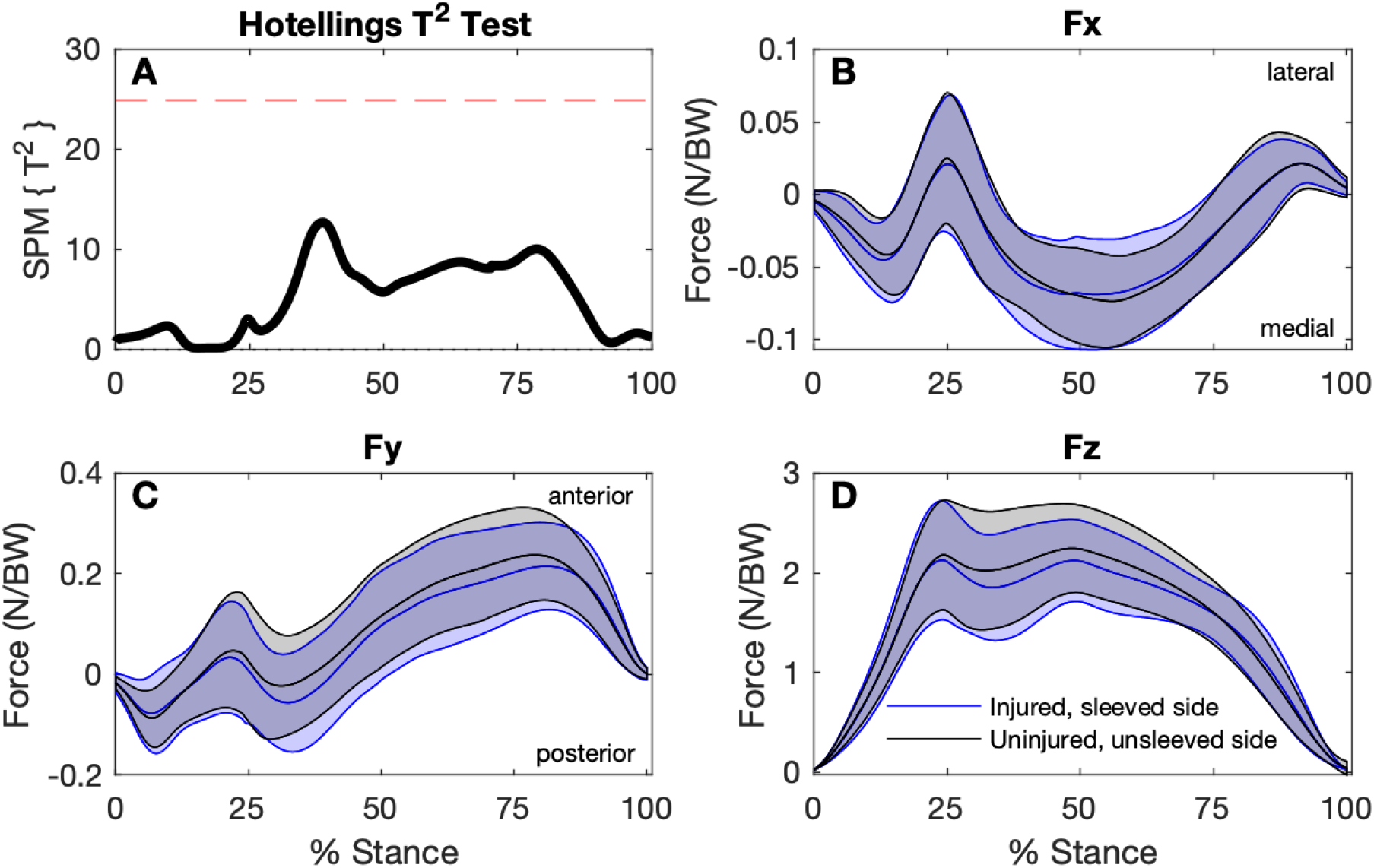
Comparison of ground reaction forces during the stance phase of the step-down hop for the ACL-injured sleeved sides to the uninjured unsleeved sides (n=30). A: SPM Hotellings T^2^ test trajectory, dashed red line indicates adjusted significance criterion; B to D: x (lateral/medial), y (anterior/posterior), and z (vertical) component curves (mean and ±1 standard deviation bands), respectively

For the discrete variables, significant effects were found for RFD, and peak anterior and posterior GRFs (Table 2). Although RFD for the sleeved, injured side was higher (34.3 ± 14.1 N/BW/s) than the unsleeved, injured side (31.9 ± 12.2 N/BW/s), this difference was not statistically significant in *post-hoc* testing. Similarly, although the actual peak FyPosterior was higher for the sleeved (0.140 ± 0.080 N/BW) than the unsleeved conditions (0.127 ± 0.071 N/BW) for the injured sides, the difference was not statistically significant in *post-hoc* testing. The (unsleeved) injured side had lower peak FyAnterior (0.237 ± 0.097 N/BW) compared to the uninjured side (0.270 ± 0.087 N/BW), and wearing the sleeve showed no statistically significant effect for the injured side.

**Table 2.**
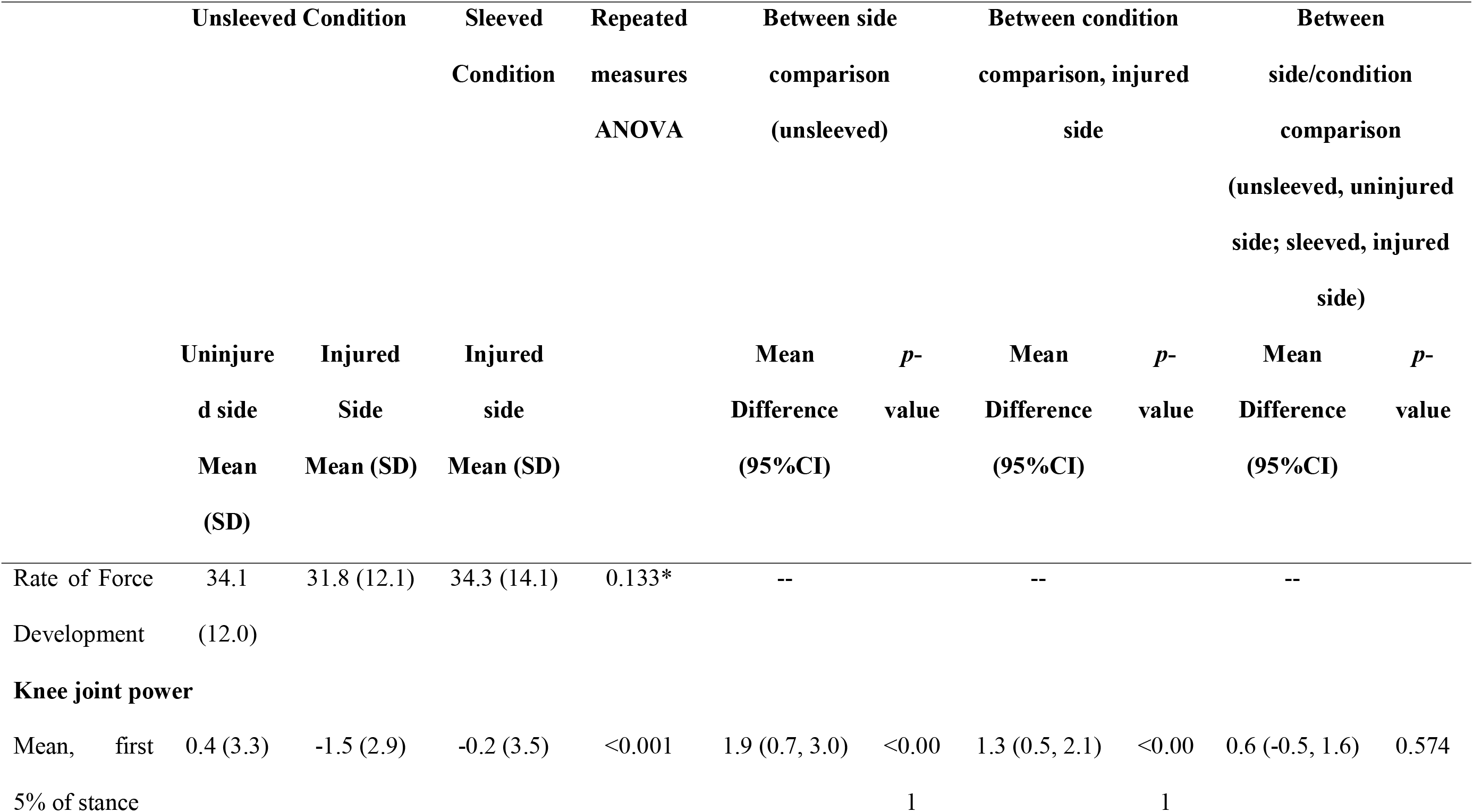
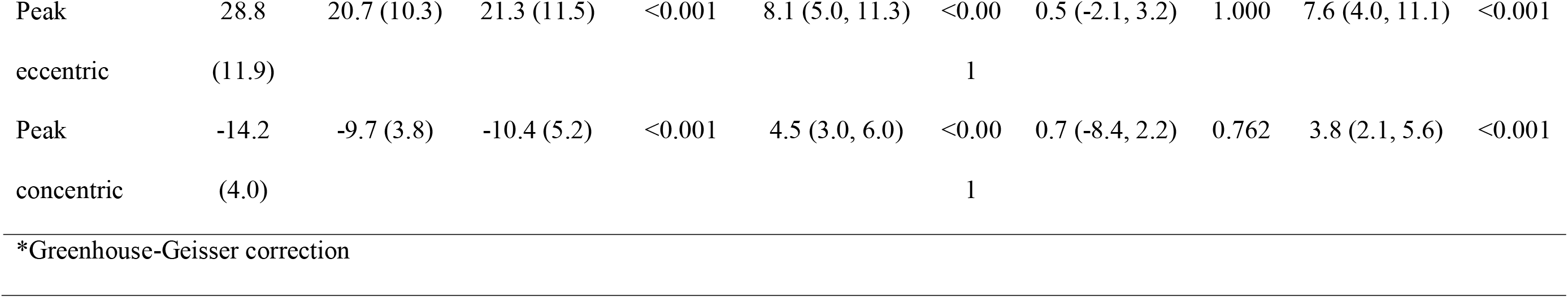
Immediate effects of wearing the sleeve: cross-over trial (n=30)

#### Knee joint power

The absorption phase (first phase) of stance entails eccentric quadriceps contraction, followed by a force generation (second) phase entailing concentric quadriceps contraction. Based on the SPM analysis, with one exception, there were no significant differences between the (unsleeved) injured and uninjured sides (Fig 5A and D), between the sleeved and unsleeved conditions for the injured side (Fig 5B and E), or between the sleeved injured and (unsleeved) uninjured sides (Fig 5C and F). The exception was at a timepoint at around 5% of the stance phase where the SPM trajectory for the comparison between the (unsleeved) injured and uninjured sides met the *p*=0.017 threshold (Fig 5A). At that timepoint, the injured side had a greater magnitude and slightly longer lasting negative power (concentric contraction) than the uninjured side. During this initial part of the landing phase there was brief a knee (external) extension moment which shifted to a flexion moment earlier for the uninjured sides. Knee angular velocity was positive for both sides during this period, indicating the knee was flexing, with the uninjured side showing a greater magnitude knee flexion velocity.

**Figure 5:**
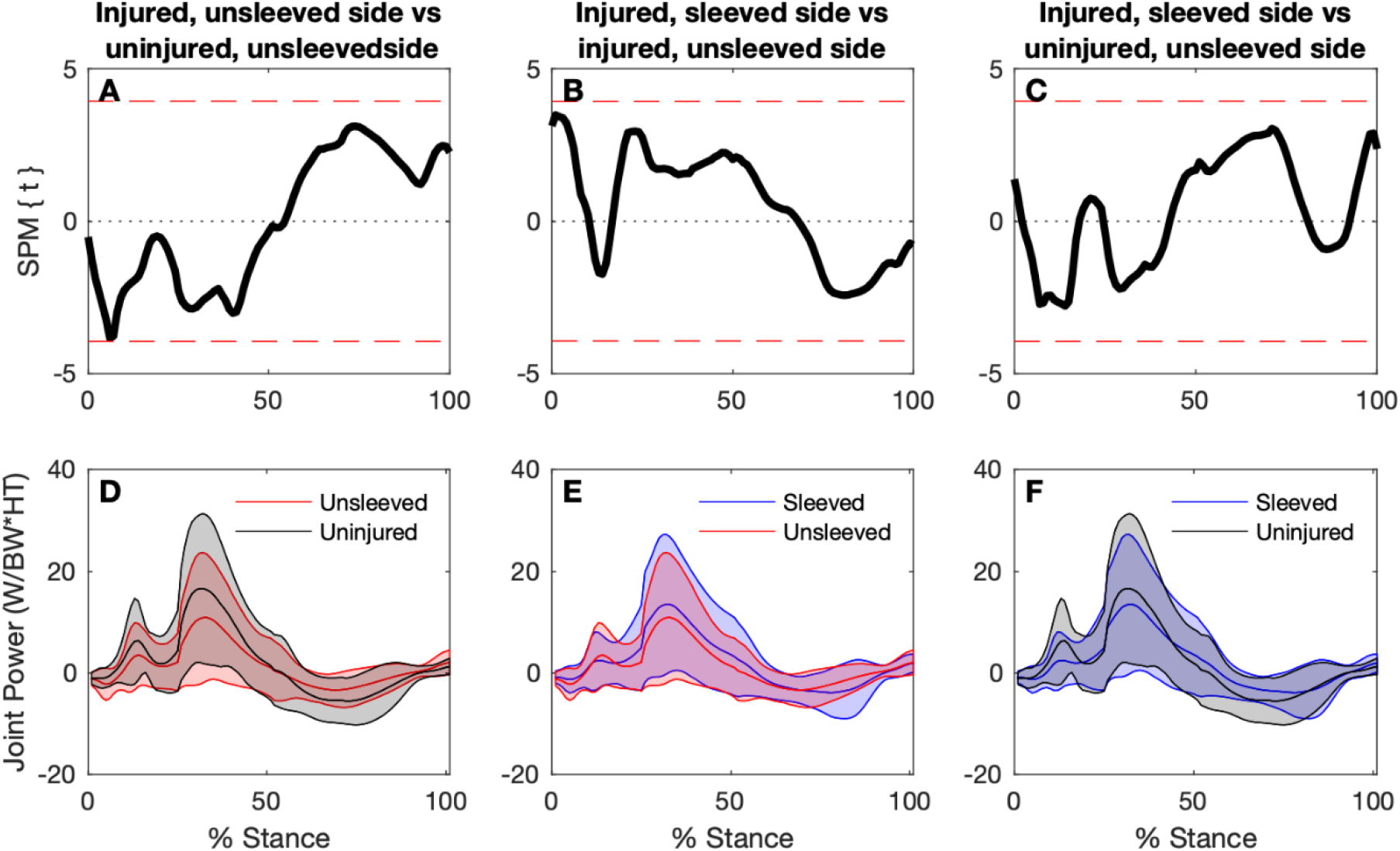
Knee joint power time-continuous comparisons for the uninjured (unsleeved) sides, and ACL injured sides for sleeved and unsleeved conditions. Top panels: SPM paired t-tests for comparisons between injured (sleeved versus unsleeved) and uninjured sides. Joint power curves (mean and ±1 standard deviation bands) for each respective test are shown in the bottom panels.

The difference for the knee power in the first 5% of the stance phase when comparing injured to uninjured side is also evident with the discrete variable analysis (Table 2). When wearing the knee sleeve, the power increased significantly for the injured side during that phase, resulting in no statistical difference when comparing the (sleeved) injured side with the (unsleeved) uninjured side. Wearing the sleeve, however, did not change the peak eccentric and concentric power, respectively (Table 2), corroborating the results of the SPM analysis.

### Part 2: Six-week effects

#### Ground reaction forces and knee joint power

There were no significant differences between the change in ground reaction force from baseline to follow-up comparing the Sleeve group to the Control group (Fig 6). Similarly, there were no significant differences in 6-week changes in joint power between groups (Fig 7).

**Fig 6.**
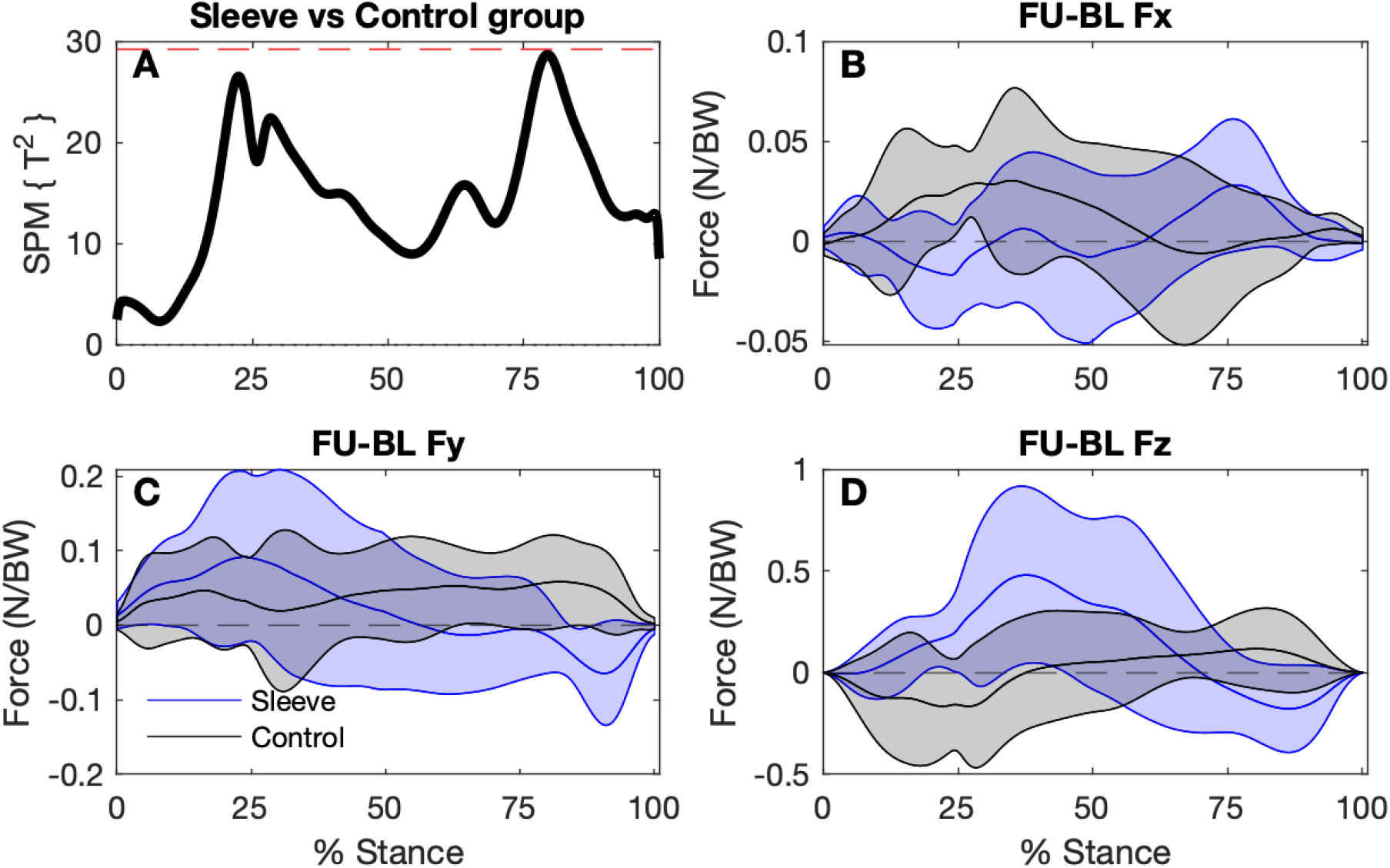
SPM analysis of ground reaction force trajectory differences between baseline and follow-up for the Sleeve group (n=9) and Control group (n=9). A. Hotellings T^2^ test trajectory. The dashed red line indicates adjusted significance criterion. B – D. The x, y, and z component curves (mean and ±1 standard deviation bands) comparing groups. Positive values indicate increases in values from baseline to follow-up.

**Fig 7.**
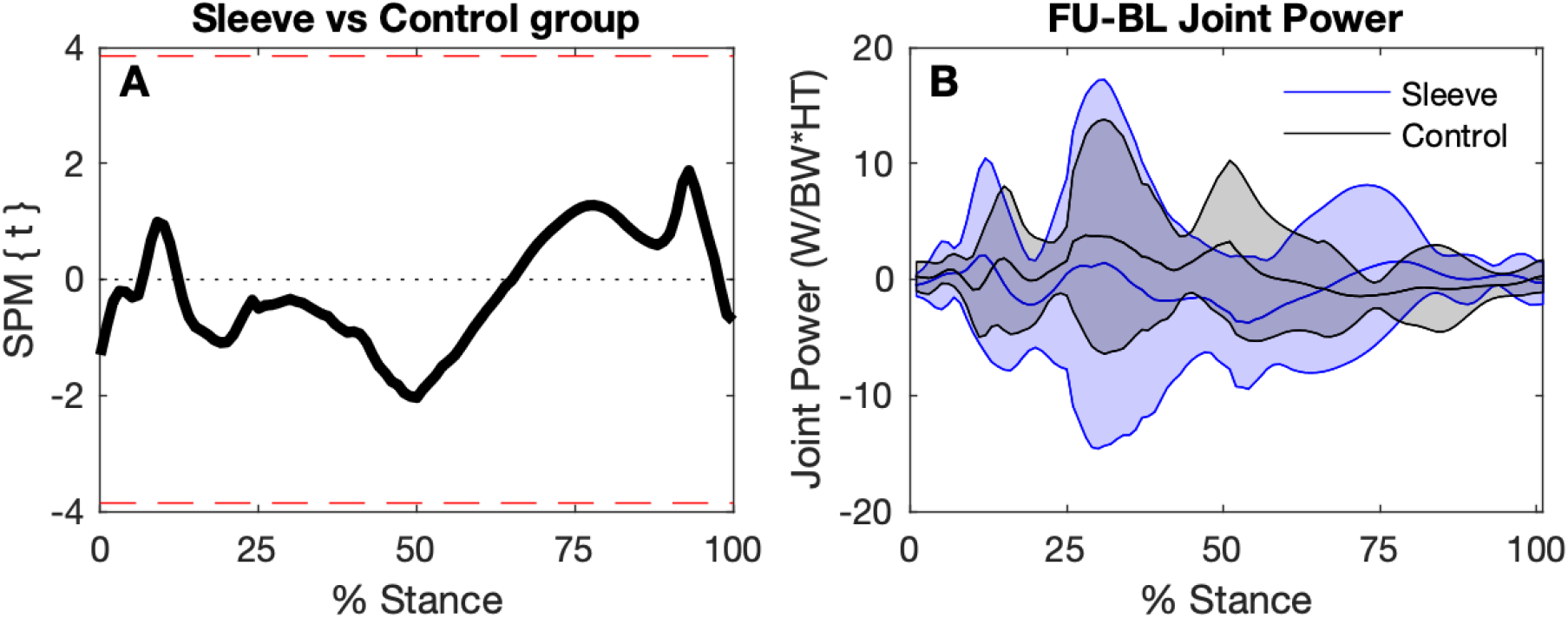
SPM comparing changes in knee joint power trajectories. A. Independent t-test trajectory (mean and ±1 standard deviation bands). Dashed red line indicates significance criterion. B. Baseline to follow-up differences for the Sleeved and the Control groups.

Discrete variable analyses of the baseline to follow-up changes suggest that there was no significant difference for the Sleeve group, based on the 95% confidence intervals for GRF and knee joint power variables (Table 3). There was, however, a difference in the response between the two groups for the peak vertical GRF (Peak FzVertical): while there was a slight *decrease* in Peak FzVertical for the Control Group, an *increase* was evident for the Sleeve Group. However, based on the 95% confidence intervals for each of the groups, the changes per group were not significant. The Control Group exhibited an increase in the anterior GRF (Peak FyAnterior) from baseline to follow-up. In contrast, no change was evident for the Sleeve group. For knee joint power in the first 5% of stance an increase was evident only for the Control group.

**Table 3.** Randomised Clinical Trial: Parameters of injured sides at baseline and follow-up, and between-group differences of changes from baseline to follow-up.

|  | Control Group (n=9) |  | Sleeve Group |  | Change score group |  | Between- |
| --- | --- | --- | --- | --- | --- | --- | --- |
|  | Mean (SD) |  | (n=9) |  | Mean difference (95% CI) |  | group |
|  |  |  | Mean (SD) |  |  |  | difference |
|  | BL | FU | BL | FU | Control | Sleeve | *p*-value* |
| Ground reaction forces |  |  |  |  |  |  |  |
| Rate of Force | 31.63 | 35.87 | 35.82 | 32.76 | -2.74 (-7.09, 1.61) | -2.11 (-7.90, 3.69) | 0.935 |
| Development | (11.82) | (11.96) | (16.24) | (10.89) |  |  |  |
| Peak FzVertical | 2.28 (0.68) | 2.32 (0.81) | 2.37 (0.56) | 2.44 (0.59) | 0.08 (-0.17, 0.33) | -0.25 (-0.79, 0.29) | 0.142 |
| Peak FxMedial | -0.07 (0.05) | -0.10 (0.05) | -0.08 (-0.04) | -0.08 (0.5) | 0.008 (-0.004, | -0.003 (-0.022, | 0.369 |
|  |  |  |  |  | 0.020) | 0.029) |  |
| Peak FxLateral | 0.07 (0.04) | 0.06 (0.04) | 0.08 (0.05) | 0.08 (0.05) | -0.014 (-0.002, | -0.003 (-0.015, | 0.086 |
|  |  |  |  |  | 0.031) | 0.010) |  |
| Peak FyAnterior | 0.23 (0.10) | 0.30 (0.10) | 0.27 (0.02) | 0.28 (0.14) | -0.064 (-0.109, - | 0.021 (-0.023, 0.065) | 0.004 |
|  |  |  |  |  | 0.019) |  |  |
| Peak FyPosterior | -0.12 (0.08) | -0.09 (0.05) | -0.14 (0.08) | -0.10 (0.07) | -0.03 (0.06) | -0.04 (-0.10, 0.01) | 0.935 |
| **Knee joint power** |  |  |  |  |  |  |  |
| Mean, first 5% of stance | -1.1 (3.5) | -1.2 (4.1) | 0.22 (4.3) | -0.38 (3.4) | -0.6 (-1.7, 0.5) | 0.2 (-2.3, 2.7) | 0.514 |
| Peak eccentric | 22.6 (12.6) | 24.7 (16.8) | 24.6 (16.3) | 26.3 (16.2) | -5.2 (-12.0, 1.5) | 1.6 (-7.3, 10.5) | 0.683 |
| Peak concentric | -12.1 (5.9) | -11.8 (6.3) | -10.7 (5.8) | -12.0 (6.1) | 1.3 (-2.2, 4.7) | -0.3 (-3.8, 3.2) | 0.221 |

BL: Baseline; FU: Follow-up; \* Mann-Whitney Test

## Discussion

In our initial report of immediate effects of wearing a knee sleeve on knee kinematics and kinetics showed increased knee flexion at initial contact and peak flexion during stance during the step-down hop [9]. In the current analysis we found, firstly, increased knee joint power during the first 5% of landing (stance) of the ACL-reconstructed knee during that task when wearing the sleeve, compared to the unsleeved condition. Wearing the knee sleeve appeared to limit an initial (external) extension moment and more quickly transition to a flexion moment, similar to the uninjured sides. Secondly, at 6-week follow-up, we found a significant difference in the *direction* of the change of the vertical GRF: while the Sleeve Group showed increased vertical GRF at follow-up, those of the Control group decreased. Per se, the differences within each group were not significant, based on the 95% confidence intervals. No other significant differences in changes between the Sleeve Group and the Control Group for kinematics and kinetics during the task were found (when not wearing the sleeve).

Our SPM results do not support our hypotheses of lower vertical GRFs, and the discrete variable analysis does not support lower RFD for the (unsleeved) injured versus the uninjured sides for the group of 30 participants. The SPM results also do not support the hypothesis that when wearing the sleeve, the peak vertical GRFs and RFD of the injured side increase significantly. However, a potential effect on RFD when wearing the sleeve may exist, although the post-hoc analyses did not reach significance.

### Ground reaction forces

Our finding of lack of immediate differences for GRF in the three planes between the injured and uninjured sides with the SPM analysis, as well as between the sleeved and unsleeved conditions contrast with previous studies that found lower vertical GRF for the ACL-reconstructed sides [20, 21]. The discrete variable analysis found significantly lower peak FyAnterior (Table 2) for the injured versus contralateral side. Peak FyAnterior occur during the early propulsion as the centre of mass is moved forwards over the weight-bearing foot. Less trunk flexion observed participants with ACL reconstruction may explain lower FyAnterior [35], but remains speculative as we did not analyse trunk movements. The knee sleeve did not influence that variable for the injured side. We used a sub-maximal hop for safety reasons as we had recruited participants who had not achieved a high level of function post-reconstruction, as defined by an IKDC less than 80/100. Dai, et al. [20] used a stop-jump task and a side-cutting task, whereas Baumgart, et al. [21] used a bilateral and a single-leg countermovement jumping task. Those tasks may have generated higher GRF than during our step-down task, thus those tasks may be able to identify residual asymmetries to a greater extent. Pietrosimone et al. [22] found that the time since ACL reconstruction influenced GRF during walking, with vertical GRF during walking gait being lower in the injured side compared to the uninjured side during the first year post-surgery, but higher than the uninjured side in the longer term.

Lower vertical GRF following ACL reconstruction may indicate a more cautious, hesitant landing pattern. Thus, the directional differences for the two groups for vertical GRF over the 6-week period may be of interest. The Sleeve Group had reported higher physical activity levels and duration during the 6-week period than the Control Group [6]. Thus, the slight *increase* in vertical GRF from baseline to follow-up for the Sleeve Group (compared to *decrease* for the Control Group), combined with the shorter stance duration [9] may reflect increased confidence as well as performance for the Sleeve Group. Such increased performance is most likely due to higher levels of physical activity during the intervention period, potentially motivated by having a sleeve available.

### Knee joint power

The stance phase of landing includes the eccentric (absorption) phase and a concentric (propulsion) phase [8, 21]. During the eccentric absorption phase, the body decelerates and the centre of mass lowers. During the concentric phase, the body is propelled upwards and forwards. Both phases have a peak FzVertical (first and second peak during the stance phase). The (unsleeved) injured side had significantly lower peak eccentric and concentric power compared to the uninjured side. Particularly during the initial 5% of stance, the unsleeved injured side had a mean concentric (negative) joint power, suggesting a greater and slightly longer lasting (external) extension moment paired with a lower magnitude (positive; flexing) angular velocity. In contrast, the uninjured sides had a mean eccentric (positive) power during that early stance phase as the flexion moment was initiated earlier. When wearing the sleeve, joint power increased during the first 5% stance for the injured side, becoming ‘more positive’, leading to similar values for the sleeved injured side and the unsleeved uninjured side. However, peak powers remained lower for the sleeved injured side compared to the uninjured side.

Our initial report suggested enhanced knee flexion angle at initial contact (mean difference 3°) when wearing the knee sleeve [9], alternatively a relatively more extended injured knee at initial contact when unsleeved. Exploring the mean and individual time series for the (external) knee flexion moments for the injured (sleeved and unsleeved) and uninjured sides (unsleeved) reveals the slightly longer (external) extension moment for the injured unsleeved sides compared to the uninjured sides in the first 5% of stance. The SPM analysis suggested that the difference at that timepoint was statistically significant [9].

### Improved sensori-motor control as a potential mechanism

Combining that result with those of the current report with reference to the slight concentric power in the early stance phase, it is possible that the unsleeved ACL-injured knee lacks knee control during landing and decreased ability to absorb initial impact. That lack of control might be evident in a short-lived extension moment and reduced angular velocity at initial contact, resulting in prolonged concentric knee power. The observed delayed flexion moment, more extended knee and reduced angular velocity at initial contact may also be explained by previously reported subtle increased quadriceps pre-activation prior to landing in a group ACL-reconstructed participants with similar duration post-surgery as our group [36]. As outlined in our earlier report [9] and by other researchers [10–12, 37, 38], wearing a knee sleeve may enhance sensori-motor control or awareness of the knee position. We speculate that wearing the sleeve might also decrease subtle fear of movement, potentially decreasing quadriceps pre-activation or guarding. Improved awareness may spontaneously lead to increased landing absorption and control, evident with slightly increased power during the first 5% of stance when sleeved. ACL ruptures are likely to occur in the first 50 ms following landing [39]. Based on our findings, we cautiously speculate that the sleeve might enhance sensori-motor mechanisms during the early eccentric (absorption) phase of landing, potentially decreasing risk for ACL injury or re-injury.

Based on our findings, changes in knee power may explain possible immediate responses of enhanced landing knee flexion when wearing the knee sleeve, rather than adaptation or responses related to ground reaction forces. To confirm such hypothesis, knee angular velocity could also be explored. In contrast, over a longer period, wearing the sleeve regularly may lead to enhanced overall performance, evident in potential small increases for vertical GRF and shorter stance duration. Whether an increased vertical GRF explains the improved stance duration for the Sleeve group at follow-up, and implications of increased anterior GRF remain speculative considering the very low sample size. Increased anterior GRF at follow-up for the Control group poses the question whether they needed greater effort to hop at the same pre-defined, individualised distance as during the baseline assessment. Furthermore, compensatory responses of the hip, ankle and trunk and centre of mass positioning during landing need to be explored further.

As a summary for the research pipeline, several directional changes are supportive of the sleeve’s role in improving function for individuals with ACL reconstruction. Immediate effects of wearing a knee sleeve included approximately 5% increased maximal single leg hop distance [6]; increased knee flexion at initial contact and peak flexion (approximately 3□) [9] and, from the current analysis, increased knee power in the first 5% of stance. While changes for RFD and peak anterior and posterior GRF when wearing the sleeve did not reach significance, directional changes were evident towards the values of the uninjured side. Particularly, the combination of increased knee flexion at initial contact and increased knee power during the early landing (stance) phase may indicate enhanced sensorimotor control during the phase in which the knee is most vulnerable for ACL rupture and re-rupture. Wearing a knee sleeve at least one hour daily for 6 weeks may lead to increased performance evident in faster stance phase during the step-down hop, and there is evidence of a directional change towards increased vertical GRF. However, there is no statistical evidence that the magnitude of GRF, peak knee power or moments changed over that period. We found no effects for wearing the sleeve in terms of self-reported outcomes (IKDC-SKF) and thigh muscle strength, indicating that wearing the sleeve did not improve nor limit self-reported knee function and muscle strength to a greater extent than not wearing the sleeve. From a clinical perspective, prescription of knee sleeves for people with ACL reconstruction should be based on assessment of the individual’s impairments, context, and their response to the knee sleeve on re-assessment.

### Methodological considerations

Our findings need to be interpreted with caution. Most variables that we explored had relatively large standard deviations compared to their means, suggesting large between-individual variability. We did not explore responses at the ankle, hip and trunk, which would add to the complexity of the analysis. Anticipation of a task (such as a drop jump) can lead to change in neuromechanical and functional differences in performance [19, 40], in turn, potentially enhancing confidence. Thus, psychological responses, such as levels of confidence, also need to be considered.

A strength of our analysis is that we included SPM as well as discrete variable analysis. SPM allows identification of differences across time series without *a priori* defined variables. However, time-aligning the respective peaks for each trial may mask potential differences between conditions and between participants. The discrete variable analysis of specified time points thus complemented the SPM analysis. As an explanatory study of findings, we performed multiple analyses, increasing the risk of Type 1 errors. On the other hand, as the COVID pandemic interfered with the follow-up sessions, only a small number of participants completed the RCT. The of Type II errors exists, thus those results, in particular, remain speculative.

Various confounders influencing the outcomes need to be considered. Wearing the knee sleeve may lead to improved knee-related confidence, or being prescribed a knee sleeve might lead to greater motivation to exercise or undertake physical activity, thereby potentially improving sensori-motor control and skill. The level of activity may influence outcomes such that those with higher performance levels may respond to a lesser degree to those with low performance (or greater impairment). We did not control for level of activity in the analysis, such as by the Tegner Activity scale. That remains a direction for future research with a larger sample. Lastly, time since surgery and sex influences biomechanical outcomes. We included participants with a large range of duration since surgery (6 months to 5 years), which may have confounded our results. We controlled for those two confounders by entering them as co-variates in the repeated measures ANOVAs of the discrete variable analyses. Thus, time since surgery and sex are unlikely to influence our findings of no statistically significant differences observed between sides for vertical GRF.

## Conclusion

Knee power increased during the first 5% of stance during a step-down hop when wearing the knee sleeve on the injured side compared to not wearing the sleeve while peak powers did not change. Wearing a knee sleeve did not change GRFs during that task for the ACL-reconstructed side, as evident in the SPM analysis. Wearing the knee sleeve at least one hour daily for 6-weeks lead to a directional change of increased vertical GRF for the Sleeve Group at follow-up. There were no significantly different changes (in magnitude) of GRFs compared to not wearing the sleeve, except for peak anterior ground reaction forces. Our previous report indicated increased knee flexion at initial contact. Combining that finding with increased knee power in the first 5% of landing when wearing the knee sleeve may indicate a protective response during the phase when the knee is most vulnerable for ACL ruptures.

## Supporting information

Supplemental Table CONSORT Checklist

## Data Availability

All relevant data are available through Zenodo at DOI: https://doi.org/10.5281/zenodo.6859069

## Abbreviations

ACL: anterior cruciate ligament
Fx: medio-lateral ground reaction force
Fy: anterior-posterior ground reaction force
Fz: vertical ground reaction force
GRF: ground reaction forces
IKDC-SKF: International Knee Documentation Committee Subjective Knee Form
RCT: randomised controlled trial
RFD: rate of force development
SPM: Statistical parametric mapping

## Acknowledgements

We thank David Jackson, Drs Anupa Patak, Sarah Ward and Mandeep Kaur for their assistance for data collection and processing.

## Ethics approval and consent to participate

Ethics approval was granted by the Health & Disability Ethics Committee, New Zealand, Reference 18/CEN/93, dated 5^th^ June 2018, amended on 27^th^ August 2019 and 4^th^ October 2019. All participants provided written informed consent to participate. All methods were undertaken in accordance with the Declaration of Helsinki.

## Authors’ contributions

Conceptualization: Gisela Sole, Niels Hammer

Data curation: Gisela Sole, Peter Lamb

Formal analysis: Gisela Sole, Peter Lamb, Todd Pataky

Funding acquisition: Gisela Sole, Niels Hammer

Investigation: Gisela Sole, Peter Lamb

Methodology: Gisela Sole, Peter Lamb, Todd Pataky

Project administration: Gisela Sole

Verification: Gisela Sole, Peter Lamb

Visualisation: Peter Lamb

Writing – original draft: Gisela Sole, Peter Lamb

Writing – review & editing: Gisela Sole, Peter Lamb, Todd Pataky, Niels Hammer

## Data availability

All relevant data are available through Zenodo at DOI: https://doi.org/10.5281/zenodo.6859069

